# Trends and Recovery Patterns of Routine Childhood Immunization Coverage in East African Community Member States, 2019-2024

**DOI:** 10.64898/2026.11.26.26369080

**Authors:** Liliane Dushimimana, Romeo Bananeza, Janvier Rukundo, Julius Niyibizi, Brian Songa, Twahiri Magoolo, Colyse Sandra Umurungi

**Affiliations:** Rwanda Food and Drugs Authority, Rwanda; University of Rwanda, Rwanda; Rwanda Implementation Science & Health Systems Lab, Kigali, Rwanda; Southern Medical University, Guangzhou, China; MSD Medipharm, Tanzania

**Keywords:** Immunization coverage, COVID-19 pandemic, post-pandemic recovery, East African Community, DTP1, DTP3, MCV1, WUENIC, dropout rate, trend analysis

## Abstract

**Background:** The COVID-19 pandemic caused unprecedented disruptions to routine childhood immunization globally, with severe consequences in low- and middle-income countries. EAC member states experienced substantial vaccination coverage setbacks during 2020–2021. This study examines immunization coverage trends and recovery across the seven EAC member states from 2019 to 2024.

**Methods:** A descriptive trend analysis was conducted using WUENIC coverage estimates for DTP1, DTP3, and MCV1 from 2019 to 2024 for the seven EAC member states, via the WHO Global Health Observatory API. An operational four-tier recovery decision aid (baseline-relative, marginal, not recovered, inconclusive) was applied using 2019 baselines, with sensitivity analysis using 2025 estimates. The framework is pragmatic, not a validated classification. DTP1-to-DTP3 dropout rates were calculated as a quality indicator.

**Results:** Recovery was highly heterogeneous. South Sudan met baseline-relative recovery criteria (all indicators ≥+2pp above baseline), though absolute coverage remained insufficient and identical 2022–2024 values may reflect non-updated estimates. Rwanda’s status was inconclusive (marginal margins reversed under 2025 data); Uganda showed marginal recovery (+1pp margins on two indicators), strengthened under 2025 data but sensitive to threshold choice. Kenya, Tanzania, Burundi, and DR Congo had not recovered by 2024. DR Congo had the steepest declines at the 2023 trough (DTP3: 73% to 60%) with widening dropout rates. Kenya declined from 2023 onward (DTP1: 99% to 93%).

**Conclusions:** Post-pandemic immunization recovery in the EAC has been uneven and incomplete. South Sudan met baseline-relative recovery criteria, though absolute coverage remained insufficient; Rwanda’s status was inconclusive and Uganda’s marginal. Persistent declines in Kenya, Tanzania, Burundi, and DR Congo underscore the need for catch-up interventions and targeted support for zero-dose children. Regional cooperation through EAC frameworks and the Big Catch-Up initiative remains critical for progress toward IA2030 targets.

## 1. Introduction

Routine childhood immunization is one of the most cost-effective public health interventions, averting an estimated 154 million deaths since the inception of the Expanded Programme on Immunization in 1974 (1,2). The COVID-19 pandemic caused the most severe disruption to global immunization services in 30 years (3,4).

Lockdowns, diversion of health personnel and resources to the pandemic response, supply chain interruptions, and public fear of visiting health facilities collectively contributed to a precipitous decline in vaccination coverage worldwide (4,5). The number of zero-dose children, those not receiving the first dose of diphtheria-tetanus-pertussis-containing vaccine (DTP1) (6), rose from 12.8 million in 2019 to a peak of 17.7 million in 2021, decreased to 13.9 million in 2022, but rose again to 14.5 million in 2023, remaining substantially above pre-pandemic levels (1,7). The WHO African Region bears the greatest burden of zero-dose children globally, accounting for 6.7 million of the 14.5 million worldwide in 2023, still above the 2019 baseline level (1,8).

The East African Community (EAC) currently comprises eight partner states. This study includes seven of them, Rwanda, Kenya, Uganda, Tanzania, Burundi, South Sudan, and the Democratic Republic of Congo, with a combined population of approximately 300 million and shared health cooperation frameworks under the EAC Treaty (9,10).

Prior to the pandemic, immunization programs in the region already faced constraints including inadequate financing, workforce shortages, and cold chain limitations (11,12). The COVID-19 pandemic exacerbated these vulnerabilities. Studies have documented substantial declines in routine immunization service delivery across African countries during the first six months of the pandemic, with 13 of 15 countries showing a decline in the monthly average number of vaccine doses provided (13), and a study from a single health post in Sierra Leone reporting a 50–80% drop in vaccinations during lockdown (14).

A systematic review estimated an overall relative median decline of 10.8% across all vaccines in low- and middle-income countries during the pandemic (15). In Eastern and Southern Africa, DTP3 coverage fell by 5 percentage points between 2019 and 2021, and 2019 marked a critical inflection point after which regional coverage shifted from an increasing to a decreasing trend (16).

In response to the pandemic-induced backsliding, global partners including WHO, UNICEF, and Gavi launched the Big Catch-Up initiative in 2023, with three strategic pillars: catching up on children missed during the pandemic years, restoring coverage to at least 2019 levels, and strengthening immunization programs within primary health care systems (3,17). The Immunization Agenda 2030 (IA2030), endorsed by the WHO African Region in 2021, provides the overarching framework for these recovery efforts, with a goal of reducing zero-dose children by 50% by 2030 (1,18).

Although Aguinaga-Ontoso et al. (16) conducted an Africa-wide joinpoint regression analysis of DTP3 coverage that included EAC countries, to our knowledge no published study has specifically evaluated post-pandemic immunization recovery across the EAC bloc. This study addresses that gap by: (i) describing DTP1, DTP3, and MCV1 coverage trends in EAC member states from 2019 to 2024; (ii) identifying the year each country returned to or surpassed its pre-pandemic (2019) coverage level; (iii) comparing DTP1-to-DTP3 dropout rates across EAC countries during the recovery period; (iv) classifying recovery status using an operational decision aid with sensitivity analyses; and (v) estimating the zero-dose burden in 2024.

## 2. Methods

### 2.1 Study Design

This study employed a descriptive trend analysis of secondary, publicly available national immunization coverage data using WUENIC data.

### 2.2 Data Source

Data were obtained from the WHO/UNICEF Estimates of National Immunization Coverage (WUENIC), 2025 revision, which provide annual country-specific coverage estimates for 195 countries and 15 vaccines (19). WUENIC estimates are derived from administrative coverage, surveys, and contextual information through an independent technical assessment by WHO and UNICEF, not formal statistical modeling, and comply with GATHER criteria (20,21).

Coverage estimates for DTP1, DTP3, and MCV1 were extracted for 2019 to 2024 via the WHO Global Health Observatory (GHO) API, using indicator codes VACCINECOVERAGE_DTP1, WHS4_100, and WHS8_110, respectively (22). The 2019 data point served as the pre-pandemic baseline, 2020-2021 as the pandemic disruption period, and 2022-2024 as the post-pandemic recovery period. Estimates for 2025 were also extracted and used only in the sensitivity analysis.

### 2.3 Study Population

The study population comprised seven of the eight current EAC partner states: Rwanda, Kenya, Uganda, Tanzania, Burundi, South Sudan, and the Democratic Republic of Congo (9). Somalia, which joined in 2024, was excluded because it had been a member for only the final year of the study period. EAC membership is used here as a descriptive grouping.

### 2.4 Variables and Indicators

Three key immunization coverage indicators were analyzed: (i) DTP1, representing access to immunization services; (ii) DTP3, representing completion of the primary DTP series and a proxy for overall program performance; and (iii) MCV1, representing measles-containing vaccine first-dose coverage. The DTP1-to-DTP3 dropout rate, calculated as [(DTP1 - DTP3) / DTP1] × 100, was used as an indicator of program quality and retention along the immunization cascade.

### 2.5 Data Analysis

Coverage trends were plotted by country and year for each indicator. Inflection points and recovery onset years were identified through systematic year-over-year comparison. This descriptive approach was chosen because the short time series (6 data points per country) precludes formal statistical modeling such as joinpoint regression (16), and is consistent with prior immunization coverage trend analyses (16,23). Non-parametric trend tests (Mann-Kendall, Sen’s slope) were considered but rejected because they test for monotonic trend, whereas the pattern of interest is non-monotonic (decline followed by rebound), and because WUENIC values are smoothed across adjacent years, violating the independence assumption underlying these tests. The study question concerns whether coverage returned to a baseline level, which is a comparison of levels rather than a test of slope.

An operational recovery decision aid was developed using a four-tier system applied to each country across all three indicators: (i) “baseline-relative recovery”: 2024 coverage ≥2pp above the 2019 baseline; (ii) “marginal recovery”: within ±1pp of baseline; (iii) “not recovered”: ≥2pp below baseline; and (iv) “inconclusive”: marginal classification that was reversed to “not recovered” under 2025 sensitivity data. A country’s overall classification was determined by its worst-performing indicator.

The ≥2pp threshold was chosen as a pragmatic margin above the 0–1pp differences that may reflect year-to-year WUENIC revisions; the marginal category acknowledges that differences of 0-1pp may not reflect genuine changes. This framework is descriptive, not statistically derived or externally validated, and the thresholds are pragmatic choices rather than guideline-based cutoffs. Its generalizability to other regions, baseline levels, or data sources remains unknown; future research should evaluate this.

The year of first recovery was identified for each country and indicator as the first year (2020–2024) in which coverage was ≥ the 2019 baseline. Recovery was classified as sustained if coverage remained ≥ baseline through 2024; if coverage later fell below baseline, the onset year was marked with an asterisk (transient return) and the overall trajectory classified as “Lost.” Countries that never reached baseline on any indicator were classified as “Never.” Where a country showed mixed patterns across indicators, the overall trajectory was classified as “Lost” if at least one indicator recovered and later fell below baseline, and as “Never” only if no indicator ever reached baseline. This onset-year analysis uses a strict ≥ baseline criterion (not the ±1pp threshold) because its purpose is to trace trajectory continuity, not endpoint status.

Two sensitivity analyses were conducted. First, 2025 WUENIC estimates were compared against 2019 baselines to determine whether including the most recent year would change any classification; countries whose marginal classification was reversed to “not recovered” were reclassified as “inconclusive”; marginal classifications that strengthened were reported but not reclassified. Second, the baseline-relative recovery threshold was varied between 1 and 3pp to test robustness to alternative cutoffs.

The estimated zero-dose burden was calculated for each EAC country as: surviving infant population × (1 − DTP1 coverage / 100), using WHO surviving infant population estimates from the World Population Prospects 2024 revision (24,25). These are approximations assuming uniform coverage distribution and may not capture geographic clustering; because coverage is reported as whole numbers, they are sensitive to rounding and are intended to convey relative scale across EAC countries, not precise counts.

All analyses were performed using Python 3.12.

## 3. Results

### 3.1 DTP1 Coverage Trends

DTP1 coverage, a measure of access to immunization services, showed diverse trajectories across EAC member states from 2019 to 2024 (Figure 1). Rwanda maintained high DTP1 coverage overall, dipping from 99% in 2019 to 94% in 2020-2021, partially recovering to 98% in 2022, dipping again to 94% in 2023, and returning to 99% by 2024. Kenya initially increased DTP1 coverage from 96% in 2019 to 99% in 2021-2022 but subsequently declined to 93% in 2024, falling below its pre-pandemic level.

**Figure 1.**
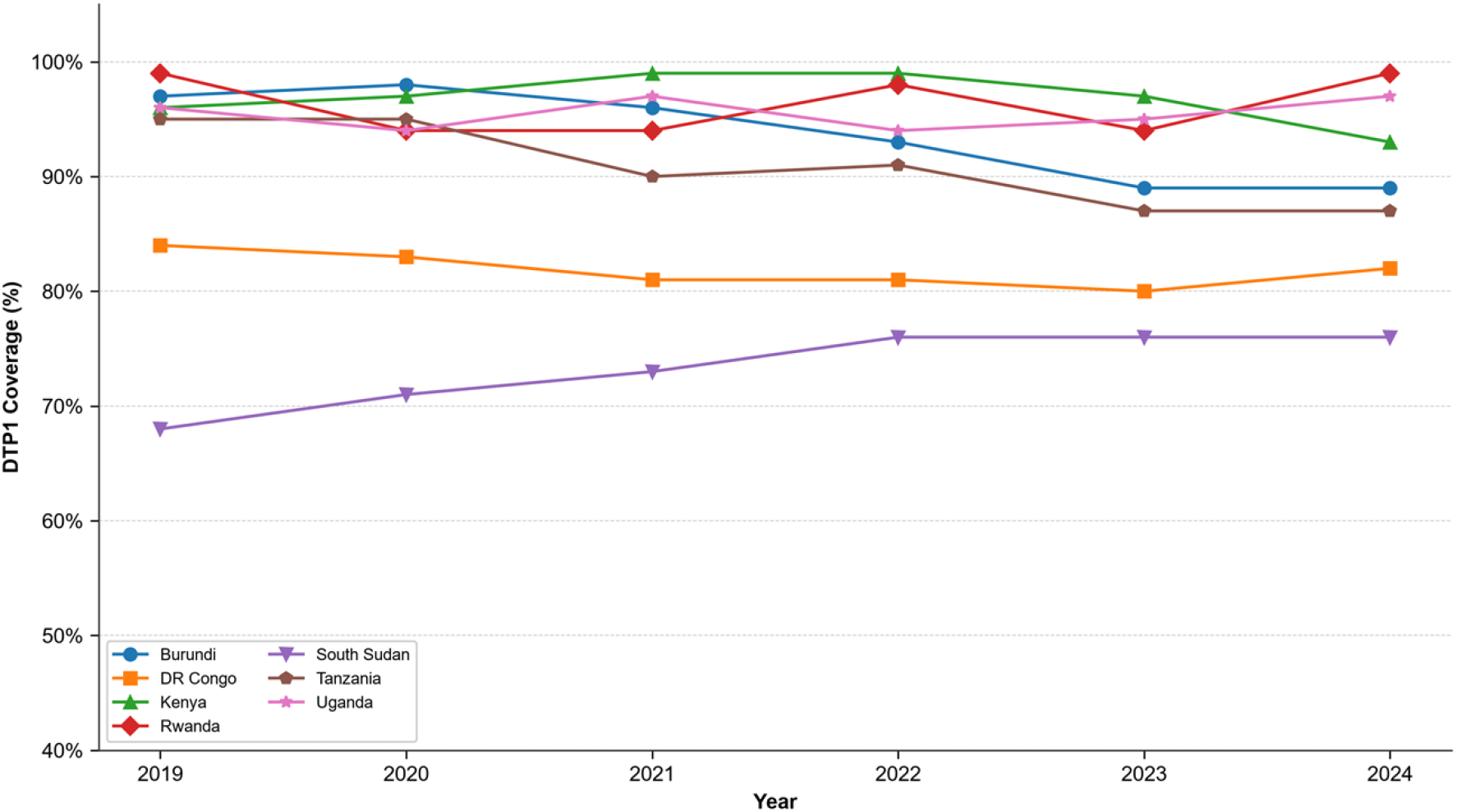
DTP1 immunization coverage trends in EAC member states, 2019-2024.

Uganda showed a resilient pattern, with coverage fluctuating between 94% and 97% and ultimately exceeding the 2019 baseline (97% in 2024 vs 96% in 2019).

Tanzania experienced an overall decline from 95% in 2019 to 87% in 2024, though the trajectory was non-monotonic: coverage fell to 90% in 2021, partially rebounded to 91% in 2022, then declined again to 87% by 2024, representing an 8 percentage-point net drop without sustained recovery. Burundi similarly declined from 97% in 2019 to 89% in 2024. South Sudan, starting from a low baseline of 68% in 2019, showed gradual improvement to 76% by 2022 but plateaued at that level through 2024. DR Congo declined from 84% in 2019 to 80% in 2023, with a slight recovery to 82% in 2024.

### 3.2 DTP3 Coverage Trends

DTP3 coverage, reflecting completion of the primary DTP vaccination series, exhibited even more pronounced disparities (Figure 2). Rwanda demonstrated a rapid recovery, dropping from 98% in 2019 to 88% in 2021 before rebounding to 98% by 2022, dipping to 94% in 2023, and returningto 98% in 2024. Uganda recovered from 89% in 2020 to 94% in 2024, surpassing the 2019 baseline of 93%. Kenya peaked at 95% in 2021 but declined to 91% in 2024, below the 2019 level of 93%.

**Figure 2.**
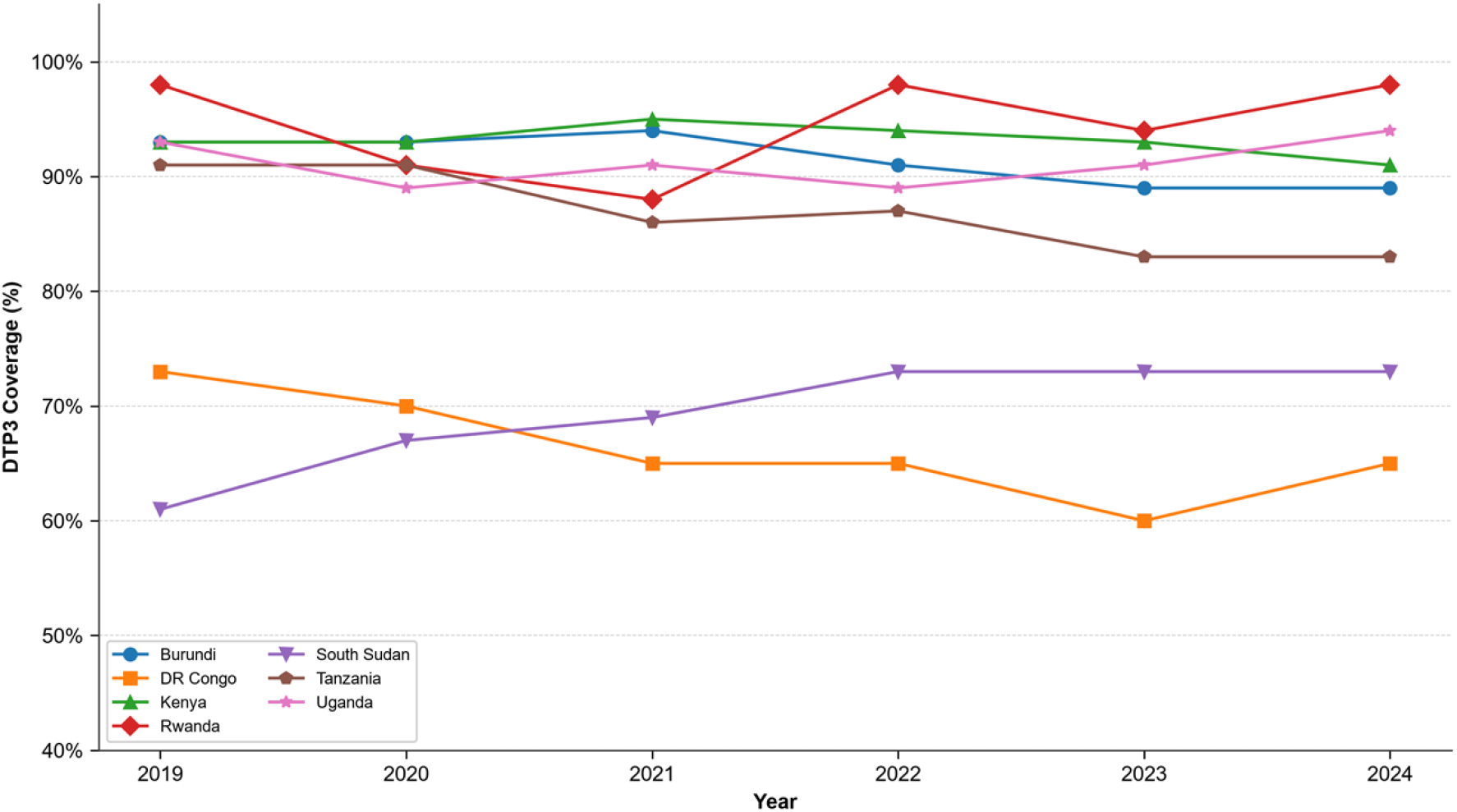
DTP3 immunization coverage trends in EAC member states, 2019-2024.

DR Congo experienced the most severe decline at the 2023 trough, with DTP3 coverage falling from 73% in 2019 to 60% in 2023, before a modest recovery to 65% in 2024, still 8 percentage points below the pre-pandemic level. Tanzania declined from 91% in 2019 to 83% in 2024. Burundi declined from 93% in 2019 to 89% in 2024. South Sudan, despite starting from the lowest baseline (61% in 2019), showed consistent improvement, surpassing its pre-pandemic level from 2020 and reaching 73% by 2022, and maintained that coverage through 2024.

### 3.3 MCV1 Coverage Trends

MCV1 coverage, indicating first-dose measles vaccination, showed variable trends across EAC countries (Figure 3). Rwanda recovered strongly, from a nadir of 87% in 2021 to a peak of 99% in 2022, before settling at 97% in 2024, still exceeding the 2019 baseline of 96%. Uganda showed steady improvement from 87% in 2019 to 92% in 2024. South Sudan improved from 65% in 2019 to a peak of 74% in 2021, before declining and plateauing at 72% from 2022 through 2024.

**Figure 3.**
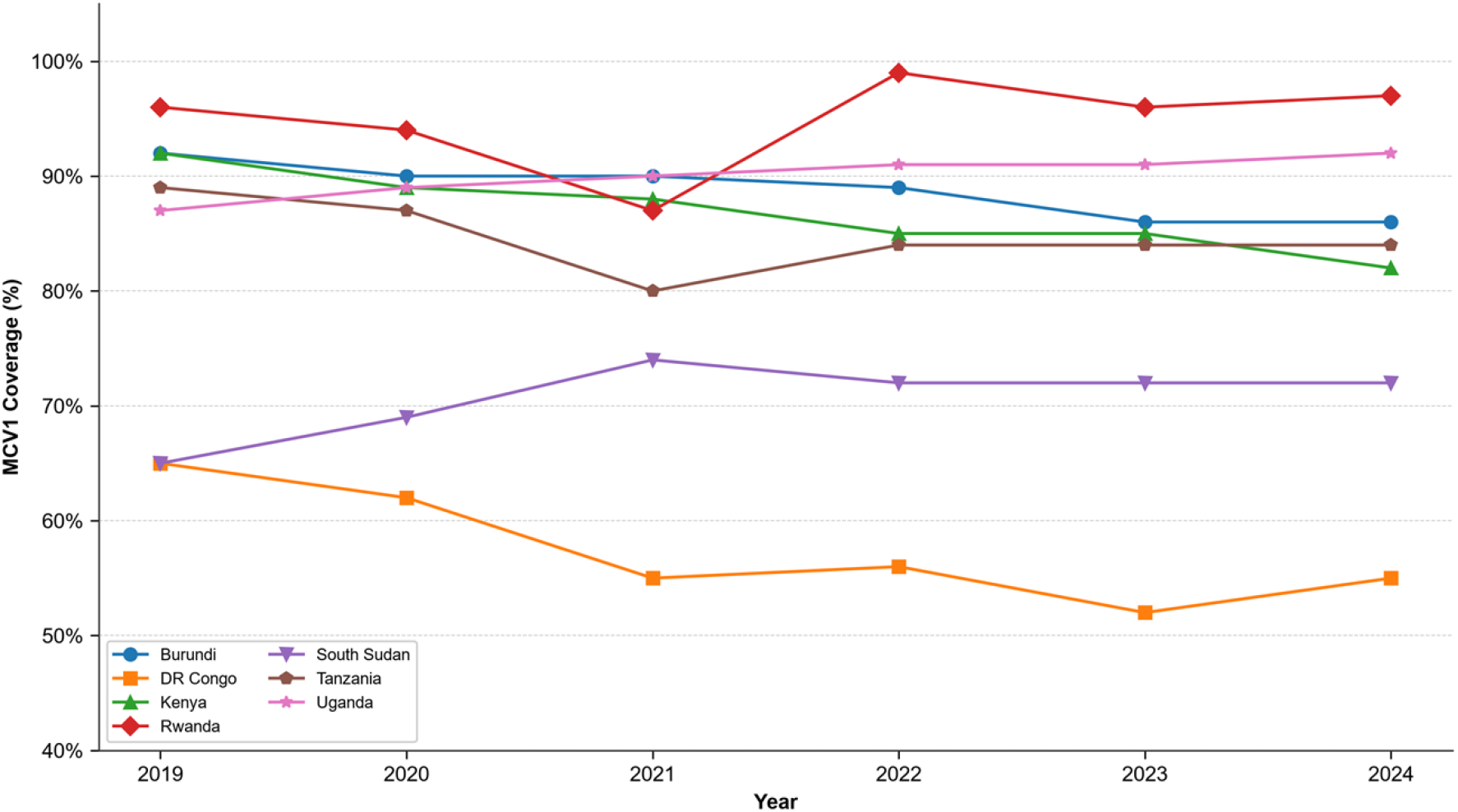
MCV1 immunization coverage trends in EAC member states, 2019-2024

Kenya experienced a persistent decline in MCV1 coverage, from 92% in 2019 to 82% in 2024, representing a 10 percentage-point drop. Tanzania declined from 89% in 2019 to a trough of 80% in 2021, with partial recovery to 84% by 2022, where coverage remained through 2024. Burundi declined from 92% in 2019 to 86% in 2024. DR Congo showed the steepest decline, from 65% in 2019 to 52% in 2023, with a slight recovery to 55% in 2024.

### 3.4 DTP1-to-DTP3 Dropout Rates

Dropout rates from DTP1 to DTP3, a key indicator of immunization program quality, varied substantially across EAC partner states and over time (Figure 4). In 2019, dropout rates were lowest in Rwanda (1.0%), Kenya (3.1%), and Uganda (3.1%), and highest in South Sudan (10.3%) and DR Congo (13.1%).

**Figure 4.**
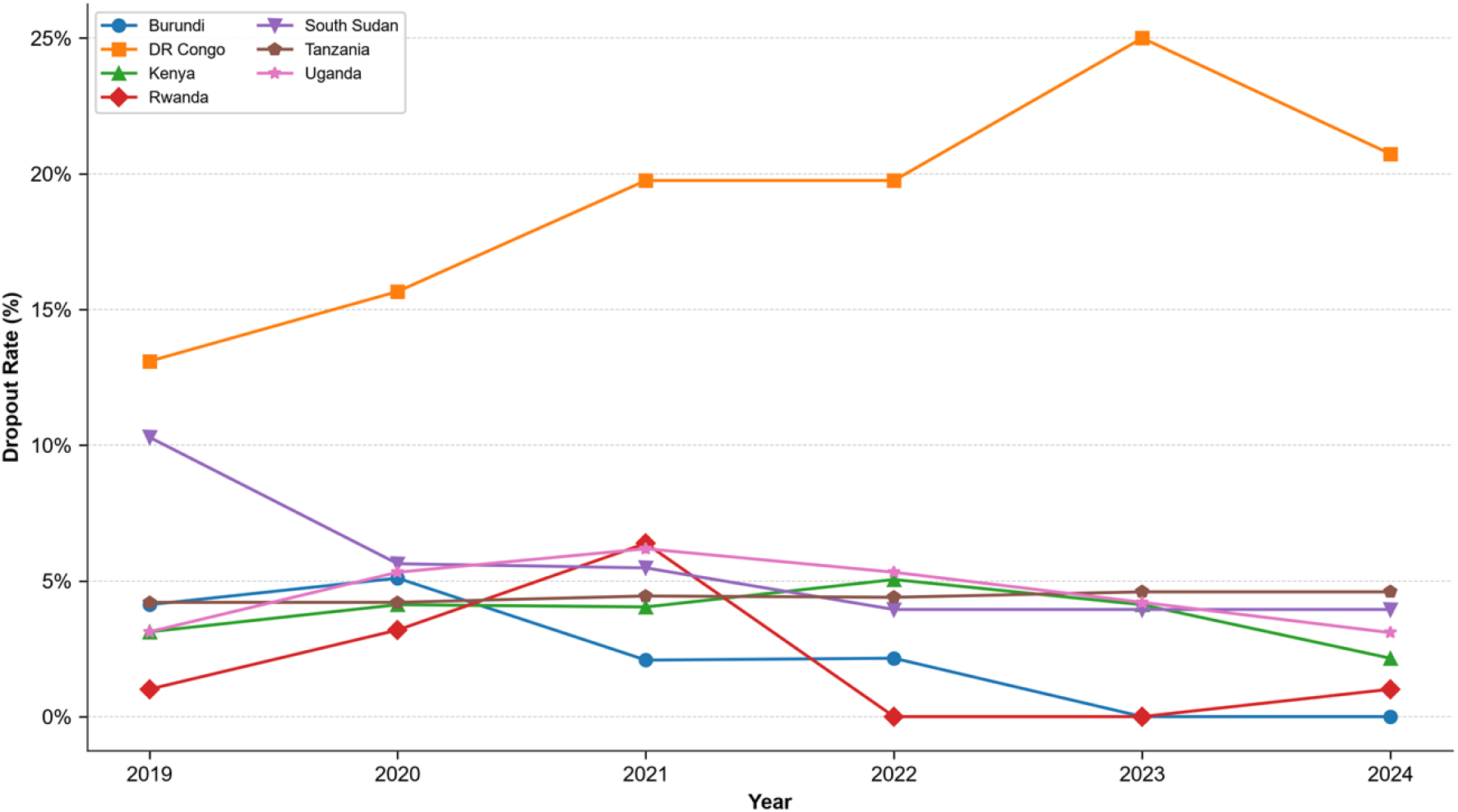
DTP1-to-DTP3 dropout rates in EAC member states, 2019-2024. Dropout rate = [(DTP1 - DTP3) / DTP1] × 100. Data source: WUENIC 2025 revision.

By 2024, Rwanda maintained a low dropout rate of 1.0%, unchanged from 2019. However, Rwanda’s dropout rate rose transiently to 3.2% in 2020 and 6.4% in 2021 during the pandemic disruption, before returning to 0% in 2022-2023. Uganda maintained its dropout rate at 3.1% in 2024, matching the 2019 level. Kenya’s dropout rate decreased from 3.1% in 2019 to 2.2% in 2024, though this occurred in the context of declining absolute coverage. Tanzania’s dropout rate remained relatively stable at approximately 4.6% in 2024 compared to 4.2% in 2019.

DR Congo showed a widening dropout rate, increasing from 13.1% in 2019 to 20.7% in 2024, consistent with challenges in retention along the vaccination cascade and an increasing proportion of children who initiate but do not complete the DTP series. South Sudan improved its dropout rate from 10.3% in 2019 to 3.9% in 2024, though this occurred in the context of low overall coverage. Burundi’s dropout rate declined from 4.1% in 2019 to 0% by 2023–2024.

### 3.5 Comparison Across Recovery Periods

Figures 5, 6, and 7 present the net percentage point change in coverage from the 2019 pre-pandemic baseline for DTP1, DTP3, and MCV1, respectively. Each figure shows the change during the pandemic period (2021 vs 2019) and the current recovery status (2024 vs 2019), with countries sorted from best to worst recovery. Positive values indicate coverage above the 2019 baseline, while negative values indicate coverage below baseline. These figures clearly illustrate the heterogeneous recovery patterns: Rwanda, South Sudan, and Uganda show positive or zero change in 2024 across all three indicators, while DR Congo, Tanzania, Burundi, and Kenya show persistent negative changes.

**Figure 5.**
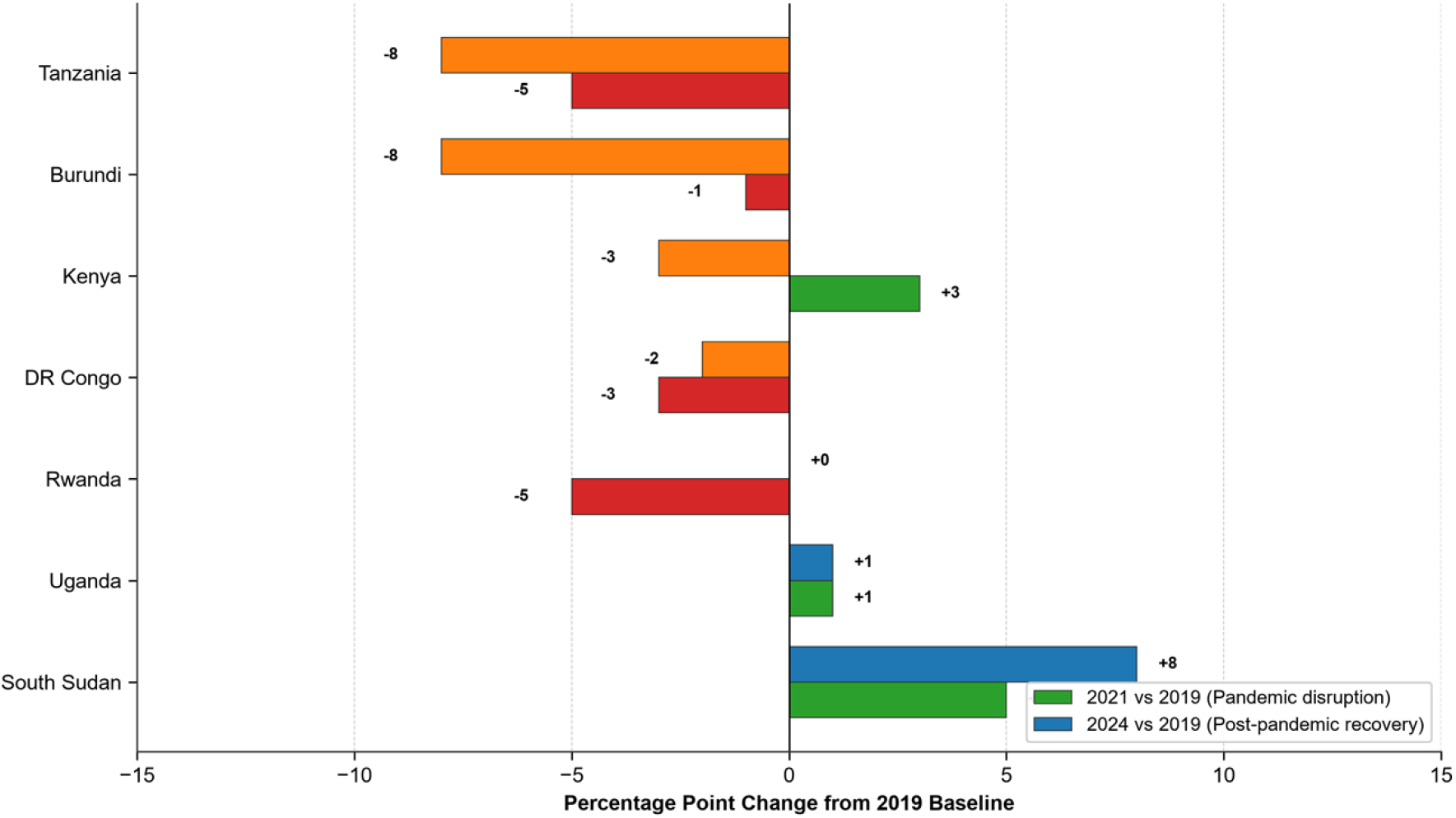
DTP1 coverage change from 2019 pre-pandemic baseline in EAC member states. Blue bars indicate improvement above baseline; orange bars indicate decline below baseline

**Figure 6.**
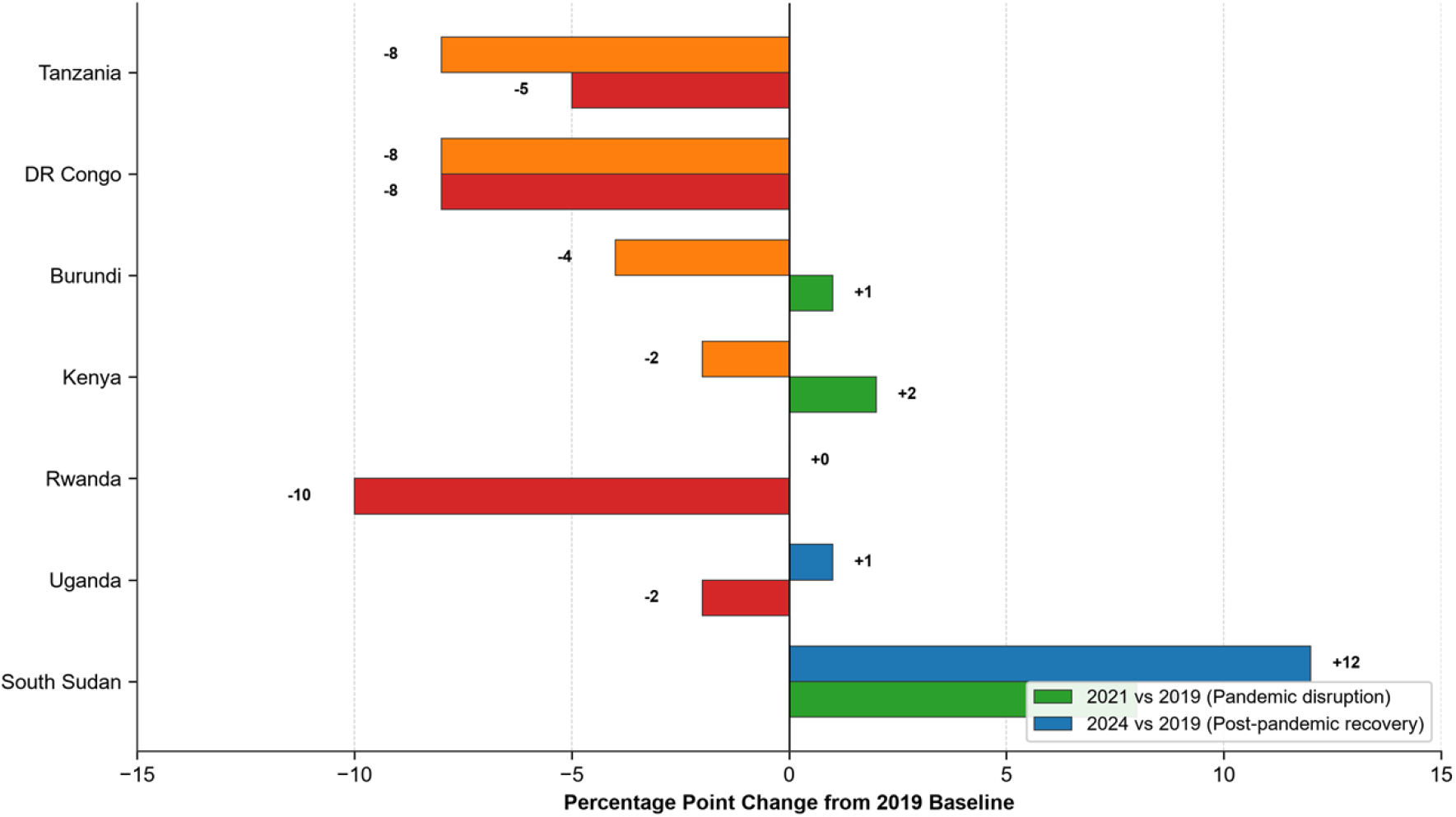
DTP3 coverage change from 2019 pre-pandemic baseline in EAC member states. Blue bars indicate improvement above baseline; orange bars indicate decline below baseline

**Figure 7.**
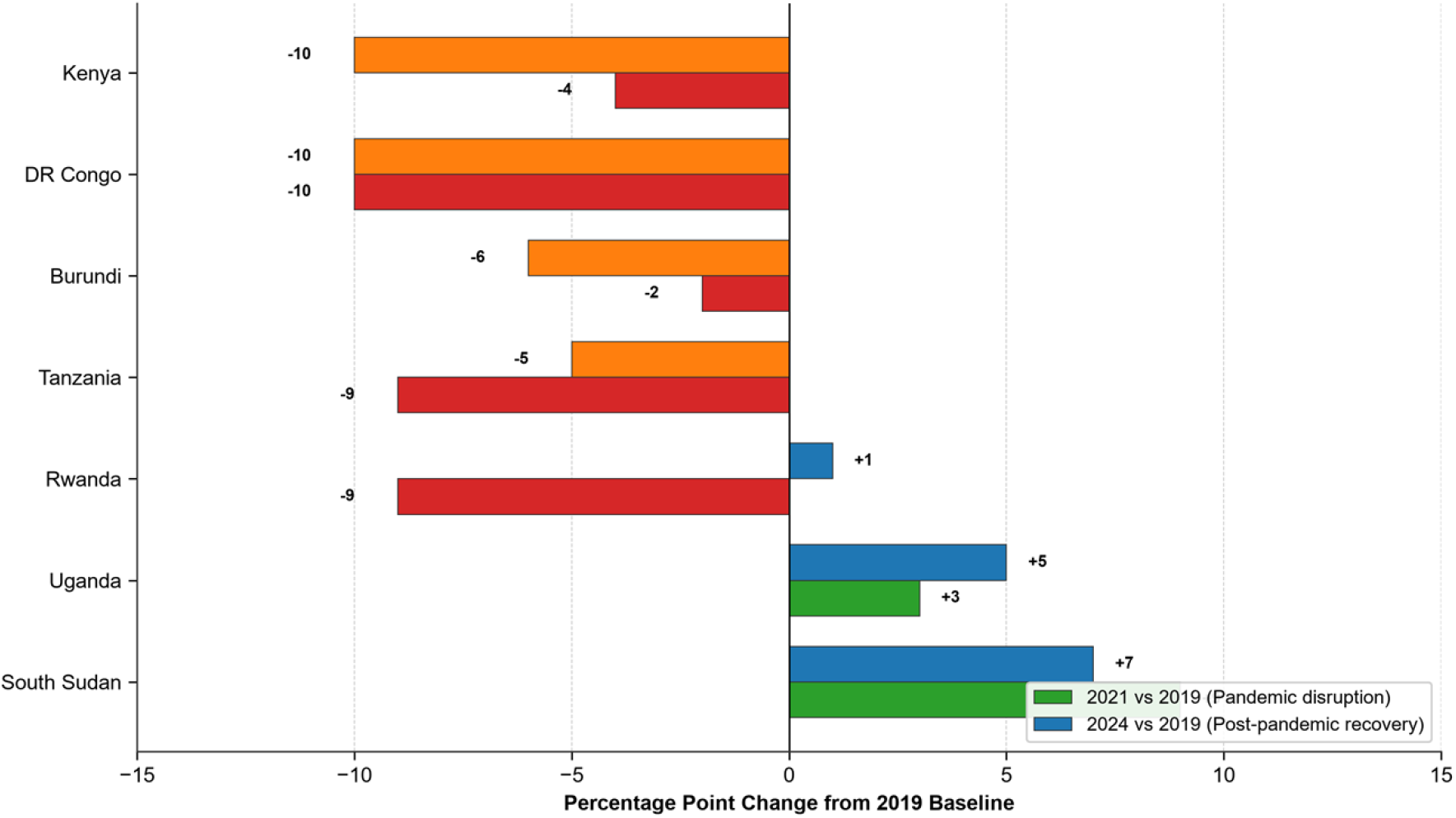
MCV1 coverage change from 2019 pre-pandemic baseline in EAC member states. Blue bars indicate improvement above baseline; orange bars indicate decline below baseline

### 3.6 Recovery Status Summary

Table 1 summarizes the recovery status of each EAC member state as of 2024, using a four-tier operational decision aid. South Sudan achieved baseline-relative recovery. Uganda showed marginal recovery. Rwanda’s classification was inconclusive: 2024 coverage met the marginal recovery criteria, but a sensitivity check using 2025 data reversed this classification to not recovered. Kenya, Tanzania, Burundi, and DR Congo had not recovered to pre-pandemic levels by 2024.

**Table 1.** Recovery status of EAC member states: 2024 coverage relative to 2019 pre-pandemic levels.

| **Country** | **DTP1 2024 (vs 2019)** | **DTP3 2024 (vs 2019)** | **MCV1 2024 (vs 2019)** | **Recovery Status** |
|---|---|---|---|---|
| Rwanda | 99% (+0) | 98% (+0) | 97% (+1) | Inconclusive |
| South Sudan | 76% (+8) | 73% (+12) | 72% (+7) | Baseline-relative |
| Uganda | 97% (+1) | 94% (+1) | 92% (+5) | Marginal |
| Kenya | 93% (-3) | 91% (-2) | 82% (-10) | Not recovered |
| Tanzania | 87% (-8) | 83% (-8) | 84% (-5) | Not recovered |
| Burundi | 89% (-8) | 89% (-4) | 86% (-6) | Not recovered |
| DR Congo | 82% (-2) | 65% (-8) | 55% (-10) | Not recovered |

### 3.7 Recovery Onset Years

The timing and sustainability of return to pre-pandemic coverage levels varied substantially across EAC member states. Using a strict criterion of coverage ≥ the exact 2019 baseline value, the first year of recovery was identified for each country and indicator, with sustained recovery defined as coverage remaining ≥ baseline through 2024 (Table 2). This trajectory analysis differs from the endpoint classification in Table 1, which uses ±1-2pp thresholds to assess recovery status as of 2024 while allowing for estimation uncertainty. A country may therefore show “marginal recovery” in Table 1 yet “Lost” in Table 2: Uganda exceeded its 2019 baseline on all three indicators by 2024 (marginal recovery) but DTP1 dipped below baseline in 2022-2023 after first reaching it in 2021 before returning in 2024 (Lost). South Sudan was the only country with a “Sustained” trajectory, where all three indicators recovered and remained above baseline from 2020 onward. Rwanda’s DTP3 and MCV1 initially recovered in 2022 but DTP3 fell below baseline in 2023 before returning in 2024; DTP1 only returned to baseline in 2024. Uganda’s MCV1 recovered earliest (2020) and was sustained, while DTP1 showed intermittent recovery before reaching baseline by 2024 and DTP3 only reached baseline in 2024. Kenya reached or surpassed baseline on DTP1 and DTP3 from 2020 through 2023 but fell below in 2024. Tanzania and Burundi briefly reached baseline on DTP1 and DTP3 in 2020 (Burundi’s DTP3 remained above baseline in 2021) but subsequently fell below and never recovered. No country recovered on MCV1 except South Sudan, Uganda, and Rwanda. DR Congo never reached its 2019 baseline on any indicator during the study period.

**Table 2.**
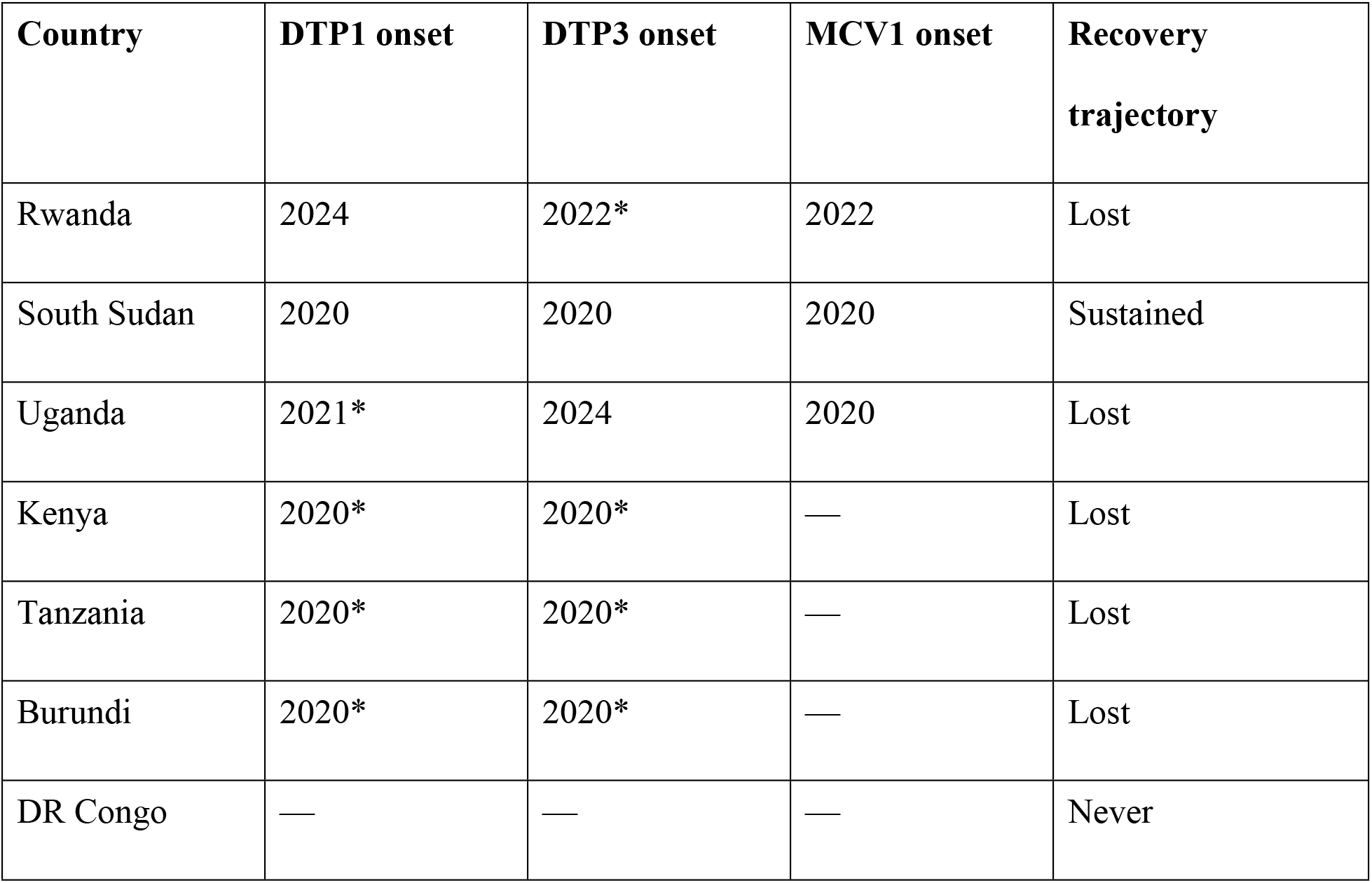
Year of first return to or surpassing 2019 pre-pandemic baseline, by country and indicator.

| **Country** | **DTP1 onset** | **DTP3 onset** | **MCV1 onset** | **Recovery trajectory** |
|---|---|---|---|---|
| Rwanda | 2024 | 2022* | 2022 | Lost |
| South Sudan | 2020 | 2020 | 2020 | Sustained |
| Uganda | 2021* | 2024 | 2020 | Lost |
| Kenya | 2020* | 2020* | — | Lost |
| Tanzania | 2020* | 2020* | — | Lost |
| Burundi | 2020* | 2020* | — | Lost |
| DR Congo | — | — | — | Never |

**Table 3.**
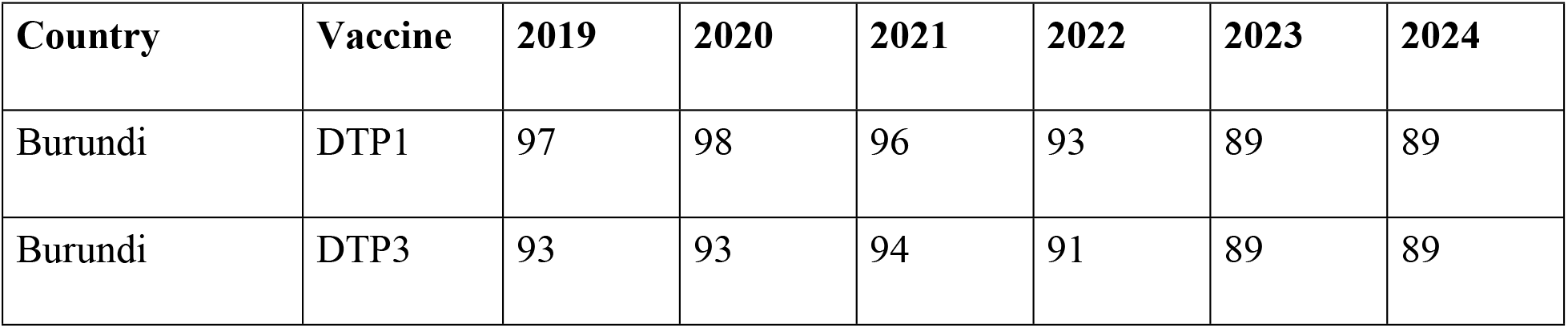
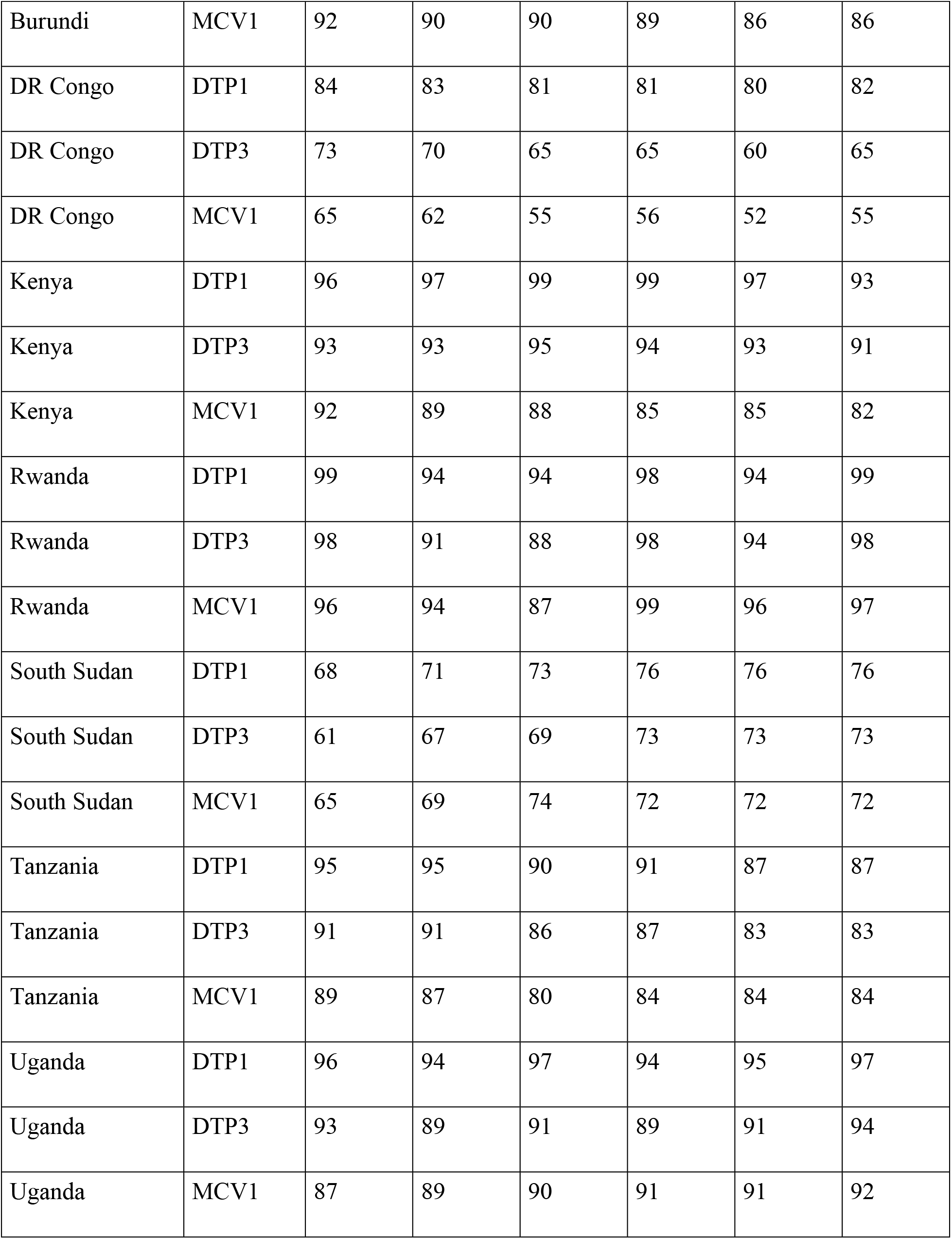
WUENIC immunization coverage estimates for EAC member states, 2019-2024.

| Country | Vaccine | 2019 | 2020 | 2021 | 2022 | 2023 | 2024 |
|---|---|---|---|---|---|---|---|
| Burundi | DTP1 | 97 | 98 | 96 | 93 | 89 | 89 |
| Burundi | DTP3 | 93 | 93 | 94 | 91 | 89 | 89 |
| Burundi | MCV1 | 92 | 90 | 90 | 89 | 86 | 86 |
| DR Congo | DTP1 | 84 | 83 | 81 | 81 | 80 | 82 |
| DR Congo | DTP3 | 73 | 70 | 65 | 65 | 60 | 65 |
| DR Congo | MCV1 | 65 | 62 | 55 | 56 | 52 | 55 |
| Kenya | DTP1 | 96 | 97 | 99 | 99 | 97 | 93 |
| Kenya | DTP3 | 93 | 93 | 95 | 94 | 93 | 91 |
| Kenya | MCV1 | 92 | 89 | 88 | 85 | 85 | 82 |
| Rwanda | DTP1 | 99 | 94 | 94 | 98 | 94 | 99 |
| Rwanda | DTP3 | 98 | 91 | 88 | 98 | 94 | 98 |
| Rwanda | MCV1 | 96 | 94 | 87 | 99 | 96 | 97 |
| South Sudan | DTP1 | 68 | 71 | 73 | 76 | 76 | 76 |
| South Sudan | DTP3 | 61 | 67 | 69 | 73 | 73 | 73 |
| South Sudan | MCV1 | 65 | 69 | 74 | 72 | 72 | 72 |
| Tanzania | DTP1 | 95 | 95 | 90 | 91 | 87 | 87 |
| Tanzania | DTP3 | 91 | 91 | 86 | 87 | 83 | 83 |
| Tanzania | MCV1 | 89 | 87 | 80 | 84 | 84 | 84 |
| Uganda | DTP1 | 96 | 94 | 97 | 94 | 95 | 97 |
| Uganda | DTP3 | 93 | 89 | 91 | 89 | 91 | 94 |
| Uganda | MCV1 | 87 | 89 | 90 | 91 | 91 | 92 |

**Table 4.** DTP1-to-DTP3 dropout rates (%) in EAC member states, 2019-2024.

| **Country** | **2019** | **2020** | **2021** | **2022** | **2023** | **2024** | **Change (2024–2019)** |
|---|---|---|---|---|---|---|---|
| Burundi | 4.1 | 5.1 | 2.1 | 2.2 | 0.0 | 0.0 | -4.1 |
| DR Congo | 13.1 | 15.7 | 19.8 | 19.8 | 25.0 | 20.7 | +7.6 |
| Kenya | 3.1 | 4.1 | 4.0 | 5.1 | 4.1 | 2.2 | -0.9 |
| Rwanda | 1.0 | 3.2 | 6.4 | 0.0 | 0.0 | 1.0 | 0.0 |
| South Sudan | 10.3 | 5.6 | 5.5 | 3.9 | 3.9 | 3.9 | -6.4 |
| Tanzania | 4.2 | 4.2 | 4.4 | 4.4 | 4.6 | 4.6 | +0.4 |
| Uganda | 3.1 | 5.3 | 6.2 | 5.3 | 4.2 | 3.1 | 0.0 |

### 3.8 Estimated Zero-Dose Children

Among EAC member states, DR Congo accounted for the largest estimated zero-dose burden in 2024 (approximately 610,000 children), followed by Tanzania (approximately 270,000), Kenya (approximately 100,000), South Sudan (approximately 85,000), Burundi (approximately 45,000), Uganda (approximately 40,000), and Rwanda (approximately 4,000). South Sudan’s zero-dose rate of 24% was the highest in the region despite its baseline-relative recovery classification. DR Congo’s burden comprised approximately 18% of its 2024 surviving infant cohort. These estimates are approximate and sensitive to rounding of reported coverage.

### 3.9 Sensitivity Analyses

Sensitivity analyses using 2025 WUENIC data reclassified Rwanda from marginal recovery to not recovered (DTP3 and MCV1 each −3 percentage points relative to 2019), while Uganda’s margins increased to ≥2 percentage points, meeting the baseline-relative threshold, and South Sudan retained its classification. Countries classified as not recovered in the primary analysis (Kenya, Tanzania, Burundi, and DR Congo) remained unchanged, although Kenya’s DTP3 deficit increased from −2 to −5 percentage points. Varying the baseline-relative recovery threshold from 1 to 3 percentage points showed that classifications were generally stable, except for Uganda, which shifted from marginal recovery to baseline-relative recovery under the 1-percentage-point threshold. These findings highlight the sensitivity of marginal classifications rather than indicating definitive changes in recovery status.

## 4. Discussion

This study examined post-pandemic recovery of routine childhood immunization coverage across the seven EAC partner states from 2019 to 2024, using WUENIC estimates from the 2025 revision. The findings reveal a highly heterogeneous recovery landscape, with South Sudan achieving baseline-relative recovery, Rwanda showing inconclusive recovery (marginal margins that reversed under 2025 data), Uganda showing marginal recovery, and Kenya, Tanzania, Burundi, and DR Congo remaining below their 2019 baselines.

Rwanda’s recovery trajectory is noteworthy, though its classification should be interpreted with caution given the inconclusive margins. DTP3 coverage dropped sharply from 98% in 2019 to 88% in 2021, rebounded to 98% in 2022, dipped again to 94% in 2023, and returned to 98% in 2024. MCV1 followed a similar volatile pattern, falling from 96% to 87% in 2021 before recovering to 99% in 2022, then settling at 97% in 2024. The DTP1-to-DTP3 dropout rate rose transiently to 6.4% in 2021 but returned to near-zero levels by 2022.

Uganda showed improvement across all three indicators, with DTP3 coverage exceeding the 2019 baseline by 2024. However, the +1pp margins on DTP1 and DTP3 are within the plausible range of year-to-year WUENIC revision changes, and the threshold sensitivity analysis confirmed that Uganda’s classification shifts from marginal to baseline-relative recovery at a 1pp cutoff. The trajectory was non-monotonic: DTP1 was below baseline in 2020, 2022 and 2023, and DTP3 from 2020 to 2023, before recovering by 2024, while MCV1 remained above baseline throughout. Notably, 2025 data strengthened Uganda’s position, with all three margins increasing to +2pp or greater, which would meet the baseline-relative threshold. Without formal uncertainty intervals or Grade of Confidence data, the distinction between marginal and baseline-relative recovery for Uganda cannot be considered robust, and the classification should be interpreted as indicative of a positive trend rather than a definitive recovery status.

South Sudan showed improvement in all three indicators despite starting from the lowest baseline. However, absolute coverage levels remain critically low (DTP3: 73% in 2024), and the classification of baseline-relative recovery should be interpreted with particular caution: all three coverage values remained identical from 2022 through 2024 (DTP1 76%, DTP3 73%, MCV1 72%), a pattern consistent with WUENIC estimates being carried forward unchanged when no new administrative or survey data are reported. This is a known feature of the WUENIC estimation process in fragile and low-capacity settings (20,26), and raises the possibility that South Sudan’s apparent recovery may partly reflect non-updated estimates rather than genuine programmatic improvement. Retrieval of Grade of Confidence ratings for these years from the WHO Immunization Data Portal would help clarify whether the flat values reflect strong empirical evidence or data gaps. Ongoing conflict, weak health infrastructure, and limited domestic financing continue to pose barriers (8,12). The dropout rate improved from 10.3% in 2019 to 3.9% in 2024, though this may partly reflect low overall coverage.

Kenya’s trajectory is concerning, with all three indicators falling below the 2019 baseline by 2024. DTP1 coverage initially rose from 96% in 2019 to 99% in 2021–2022, then declined to 97% in 2023 and further to 93% in 2024. DTP3 followed a similar pattern, peaking at 95% in 2021 before declining to 91% in 2024, 2 percentage points below baseline. MCV1 showed an overall declining trend from 92% in 2019 to 82% in 2024, with the decline pausing at 85% in 2022–2023 before resuming in 2024. The 2025 sensitivity check confirmed further deterioration, with DTP3 declining to 88% (–5pp from baseline) and all three indicators worsening. WUENIC estimates for Kenya are challenged by reported administrative data: the 2025 WUENIC DTP3 estimate of 88% exceeds reported coverage of 80%, and WHO and UNICEF note that an ongoing 2025 Interim DHS survey may revise the time series (19). Despite these coverage declines, the DTP1-to-DTP3 dropout rate improved from 5.1% in 2022 to 2.2% in 2024, suggesting some progress in follow-up completion among children who received DTP1. The WHO African Region IA2030 progress report concluded that coverage remains below targets and equity gaps persist across the region (8).

Tanzania and Burundi showed net declines across all three indicators without baseline-relative recovery by 2024. Tanzania’s DTP3 fell from 91% to 83% (–8pp), DTP1 from 95% to 87% (– 8pp), and MCV1 from 89% to 84% (–5pp); Burundi’s DTP3 fell from 93% to 89% (–4pp), DTP1 from 97% to 89% (–8pp), and MCV1 from 92% to 86% (–6pp). Tanzania’s trajectory was non-monotonic, with a partial recovery in 2022, while Burundi’s declines were more sustained. All six values remained flat from 2023 through 2025, consistent with carried-forward estimates (20,26); WUENIC notes confirm Tanzania’s 2025 DTP3 estimate was calibrated to 2023 levels with reported data excluded (19). Aguinaga-Ontoso et al. (16) identified Tanzania among 26 African countries with DTP3 declines between 2019 and 2021, though those estimates (89%, 81%) have since been revised upward; Burundi showed no decline in that analysis. Despite overall coverage losses, Burundi’s dropout rate improved from 4.1% to 0.0%.

DR Congo was the only EAC member state that never reached baseline on any indicator during 2020-2024. DTP3 fell from 73% to 60% in 2023, recovering to 65% in 2024 (−8pp); DTP1 declined from 84% to 82% (−2pp) and MCV1 from 65% to 55% (−10pp). The dropout rate peaked at 25.0% in 2023 before improving to 20.7% in 2024. All three indicators remained flat from 2024 to 2025. DR Congo is among the ten countries accounting for 80.3% of zero-dose children in the WHO African Region during 2019-2022 (27). Ongoing conflict and weak health system infrastructure contribute to these challenges (12,28); WUENIC notes indicate reported data were excluded and the DTP3 estimate was informed by extrapolation, with an ongoing survey that may revise the series (19). Disentangling the pandemic’s contribution from pre-existing structural barriers is not possible with available data.

The heterogeneous recovery patterns are consistent with global assessments. The IA2030 Global Progress Report 2024 noted that initial post-pandemic recovery has not been sustained globally, though some progress was seen in the WHO African Region (1). The WHO Executive Board report highlighted that recovery has been uneven, with least progress in low-income countries (7). This study is partially consistent: while DR Congo and Burundi experienced minimal recovery, South Sudan, also low-income, achieved baseline-relative recovery from a low baseline, suggesting that income status alone does not determine outcomes.

The observed differences in recovery trajectories across EAC countries may reflect multiple factors, including health system governance, adaptation of service delivery during and after the pandemic, disruption to integrated supportive supervision (29), catch-up campaign intensity (3,17), and the extent of pandemic-related disruption to routine immunization services, including health worker redeployment for COVID-19 response (13) and reduced service utilization due to fear of contagion and movement restrictions (13,14). However, this descriptive study cannot establish causal relationships between any specific factor and observed coverage changes. These potential explanations should be considered hypotheses for future investigation rather than evidence-based explanations.

The Big Catch-Up initiative, launched in 2023 by WHO, UNICEF, and Gavi, represents a critical intervention for addressing the immunity gaps identified in this study (3,17). Its three pillars of catch-up, restore, and strengthen align directly with EAC needs. However, activities only began in 2024 (1), which may explain why recovery remains incomplete. The full impact may not be reflected in 2024 estimates and may become evident in future years.

### 4.1 Strengths and Limitations

This study uses the most recent WUENIC estimates (2025 revision), includes the seven EAC partner states, and evaluates three key childhood immunization indicators (DTP1, DTP3, and MCV1) together with DTP1-to-DTP3 dropout rates. To our knowledge, this is the first study to specifically assess post-pandemic routine immunization recovery across the EAC during the 2019– 2024 period.

Several limitations should be considered. First, WUENIC estimates depend on the availability and quality of underlying empirical data, which differ across countries and over time (20,26). Estimates may be carried forward when new empirical data are unavailable; for example, South Sudan reported identical values for all three indicators between 2022 and 2024, DR Congo’s DTP1 and DTP3 remained unchanged between 2021 and 2022, and Burundi’s DTP1 and DTP3 were identical in 2023 and 2024. Such stability may represent genuine consistency or may reflect limited data availability. Grade of Confidence ratings, which could provide additional information on estimate reliability, were not incorporated because they are not available through the WHO Global Health Observatory API (21).

Second, the six-year observation period provides limited data points for each country and does not support formal time-series modeling; therefore, findings should be interpreted as exploratory trend descriptions rather than confirmatory analyses. Third, national-level estimates may conceal substantial subnational variation, particularly in geographically large and heterogeneous countries such as DR Congo and Tanzania. Future research should incorporate district-level data and equity dimensions to better characterize disparities. Fourth, the descriptive design prevents causal inference regarding the factors or interventions contributing to observed recovery patterns. Fifth, reliance on a single data source without triangulation against independent survey sources, such as DHS or MICS, may limit robustness, particularly for marginal classifications determined by small (0–1 percentage point) differences.

Sixth, the four-tier recovery framework developed in this study is an investigator-defined operational approach based on heuristic thresholds and has not undergone formal validation. Classifications for Rwanda (0 percentage-point margins on two indicators) and Uganda (1 percentage-point margins on two indicators) should therefore be interpreted cautiously, whereas South Sudan’s classification was more robust, with all recovery margins ≥7 percentage points. The applicability of this framework to other regions or datasets remains uncertain. Nevertheless, comparison with WHO regional assessments showed broad consistency: DR Congo and Burundi continue to be identified as countries facing persistent immunization challenges (8), while South Sudan’s improvement aligns with documented gradual expansion of immunization services (27), providing limited external support for the observed patterns. Future WUENIC revisions may also modify historical estimates and alter recovery classifications.

This study was reported according to the STROBE guidelines for observational studies using secondary data (30) and considered relevant GATHER criteria where applicable (31).

### 4.2 Implications for Policy and Practice

The findings of this study have several important implications for EAC health ministries and immunization partners. First, the persistent coverage declines observed in DR Congo, Tanzania, Burundi, and Kenya highlight the need for intensified catch-up vaccination efforts, particularly among zero-dose and under-vaccinated children who have accumulated since the COVID-19 pandemic. The Big Catch-Up initiative provides an appropriate framework for these activities (3,17), but effective implementation will require sustained political commitment, adequate financing, and strengthened health system capacity.

Second, the EAC regional cooperation framework offers opportunities for cross-country learning and technical collaboration. Although Rwanda’s recovery status was inconclusive and remains sensitive to subsequent WUENIC revisions, its comparatively favorable trajectory through 2024 warrants further investigation to identify potentially transferable programmatic practices. The EAC Centre of Excellence for Vaccine, Immunization and Health Supply Chain Management, established with support from the German Development Bank (KfW) and hosted at the University of Rwanda (32), provides an important platform for regional knowledge exchange, capacity building, and dissemination of best practices.

Third, declining MCV1 coverage in Kenya, Tanzania, Burundi, and DR Congo is of particular concern because it increases the risk of measles outbreaks. The WHO African Region reported 66 large outbreaks of vaccine-preventable diseases in 2023, including multiple measles outbreaks (8), while pandemic-related disruptions to measles surveillance further increased the likelihood of delayed outbreak detection (33). Strengthening measles surveillance, restoring high routine MCV1 coverage, and implementing targeted supplementary immunization activities should therefore remain public health priorities.

Fourth, recovery strategies should prioritize equity by focusing on rural, conflict-affected, and displaced populations, which account for a disproportionate share of zero-dose and under-vaccinated children (6). Strengthened subnational micro-planning, routine district-level coverage monitoring, and the use of equity stratifiers are needed to ensure that improvements in national coverage are reflected across underserved populations rather than masking persistent geographic disparities.

Finally, sustained political commitment and increased domestic investment in immunization are essential to consolidate recovery and accelerate progress toward Immunization Agenda 2030 (IA2030) targets. The finding that only 16 of 47 countries in the WHO African Region achieved the 90% DTP3 coverage target in 2023 (8) illustrates that immunization recovery remains a regional challenge requiring long-term investment in resilient routine immunization systems rather than short-term recovery efforts alone.

## 5. Conclusion

Post-pandemic recovery of routine childhood immunization coverage in the EAC has been uneven and incomplete as of 2024. Under the operational recovery framework used in this study, South Sudan met the predefined criterion for baseline-relative recovery, but absolute coverage levels remain critically low (DTP3: 73% in 2024) and identical 2022–2024 estimates may partly reflect non-updated WUENIC values rather than genuine programmatic improvement. Rwanda’s status was inconclusive (margins that reversed under 2025 data), and Uganda showed marginal recovery based on +1pp margins sensitive to threshold choice. Kenya, Tanzania, Burundi, and DR Congo remain below their 2019 baselines, with DR Congo experiencing the most severe decline at the 2023 trough and a dropout rate that widened through 2023 before partial improvement in 2024, suggesting challenges in retention along the vaccination cascade that warrant monitoring. The Big Catch-Up initiative provides a critical framework for recovery, but its implementation must be accelerated. Regional cooperation through EAC frameworks, sustained political commitment, and targeted support for zero-dose children are essential for achieving IA2030 goals.

## Ethics Statement

This study was a secondary analysis of publicly available, aggregated national-level immunization coverage estimates from the WHO/UNICEF Estimates of National Immunization Coverage (WUENIC). It involved no human participants, no individual-level data, and no personally identifiable information. Ethical approval and informed consent were therefore not required.

## Funding

The authors received no specific funding for this work.

## Competing Interests

The authors have declared that no competing interests exist.

## Data Availability Statement

All data used in this study are publicly available from the WHO/UNICEF Estimates of National Immunization Coverage (WUENIC) database, accessible through the WHO Immunization Data Portal (https://immunizationdata.who.int/) and the WHO Global Health Observatory API (https://ghoapi.azureedge.net/api/). The analysis codes are available in the EAC Immunization Recovery Analysis Code repository at https://doi.org/10.5281/zenodo.24464703 *Data were accessed on 20 July 2026*.

## Author Contributions

Conceptualization: LD, RB, JN. Data curation: RB. Formal analysis: RB. Investigation: RB. Methodology: RB, LD, JN. Project administration: RB. Software: RB. Supervision: JN. Validation: LD, JR, BS. Visualization: RB. Writing – original draft: RB. Writing – review & editing: LD, RB, JR, JN, BS, TM, CSU.

## Notes

### Competing Interest Statement

The authors have declared no competing interest.

## References

1. Immunization Agenda 2030. IA2030 Global Progress Report 2024. Geneva: WHO; 2024. Available from: https://www.immunizationagenda2030.org/images/documents/Immunization_Agenda_2030_Global_Progress_Report_2024_final.pdf

2. Shattock AJ, Johnson HC, yoon Sim S, et al. Contribution of vaccination to improved survival and health: modelling 50 years of the Expanded Programme on Immunization. The Lancet. 2024;403(10441):2307–2316. doi:10.1016/S0140-6736(24)00850-X

3. World Health Organization, UNICEF, Gavi. An Essential Immunization Recovery Plan: Catch-Up, Restore and Strengthen. Geneva: WHO; 2023. Available from: https://www.immunizationagenda2030.org/images/documents/9789240075511-eng.pdf

4. Lassi ZS, Naseem R, Salam RA, Siddiqui F, Das JK. The Impact of the COVID-19 Pandemic on Immunization Campaigns and Programs: A Systematic Review. International Journal of Environmental Research and Public Health. 2021;18(3):988. doi:10.3390/ijerph18030988

5. Shapira G, Ahmed T, Drouard SHP, Fernandez PA, Kandpal E, Nzelu C, et al. Disruptions in maternal and child health service utilization during COVID-19: analysis from eight sub-Saharan African countries. Health Policy and Planning. 2021;36(7):1140–1151. doi:10.1093/heapol/czab064

6. United Nations Children’s Fund. The State of the World’s Children 2023: For Every Child, Vaccination. New York: UNICEF; 2023. Available from: https://www.unicef.org/reports/state-worlds-children-2023

7. World Health Organization. Progress towards global immunization goals and implementation of the Immunization Agenda 2030. Geneva: WHO; 2024. Document B154/9. Available from: https://apps.who.int/gb/ebwha/pdf_files/EB154/B154_9-en.pdf

8. Bekele AT, Masresha BG, Bita Fouda AA, Mitula P, Bwaka A, Wanyoike S, et al. Progress on implementation of immunization agenda 2030 (IA2030) in the WHO African region: achievements, lessons learned and priorities. Pan African Medical Journal. 2025;51(Suppl):48959. doi:10.11604/pamj.supp.2025.51.1.48959

9. East African Community. Treaty for the Establishment of the East African Community. Arusha: EAC; 1999. Available from: https://www.eac.int/documents/category/treaties

10. East African Community. EAC Medicines and Health Technologies Policy. Arusha: EAC; 2018. Available from: https://www.eac.int/documents

11. Mihigo R, Okeibunor J, Anya B, Mkanda P, Zawaira F. Challenges of immunization in the African Region. Pan African Medical Journal. 2017;27(Suppl 3):12. doi:10.11604/pamj.supp.2017.27.3.12127

12. Nshimirimana D, Mihigo R, Clements CJ. Routine immunization services in Africa: back to basics. Journal of Vaccines & Immunization. 2013;1(1):6–12. doi:10.14312/2053-1273.2013-2

13. Masresha B, Luce R, Shibeshi M, Ntsama B, N’Diaye A, Chakauya J, et al. The performance of routine immunization in selected African countries during the first six months of the COVID-19 pandemic. Pan African Medical Journal. 2020;37(Suppl 1):12. doi:10.11604/pamj.supp.2020.37.1.26107

14. Buonsenso D, Cinicola B, Kallon MN, Iodice F. Child Healthcare and Immunizations in Sub-Saharan Africa During the COVID-19 Pandemic. Frontiers in Pediatrics. 2020;8:517. doi:10.3389/fped.2020.00517

15. Cardoso Pinto AM, Ranasinghe L, Dodd PJ, Budhathoki SS, Seddon JA, Whittaker E. Disruptions to routine childhood vaccinations in low- and middle-income countries during the COVID-19 pandemic: A systematic review. Frontiers in Pediatrics. 2022;10:979769. doi:10.3389/fped.2022.979769

16. Aguinaga-Ontoso I, Guillen-Aguinaga S, Guillen-Aguinaga L, Alas-Brun R, Onambele L, Aguinaga-Ontoso E, et al. COVID-19 Impact on DTP Vaccination Trends in Africa: A Joinpoint Regression Analysis. Vaccines. 2023;11(6):1103. doi:10.3390/vaccines11061103

17. World Health Organization Regional Office for Africa. Progress Report on the Framework for the Implementation of the Immunization Agenda 2030 in the WHO African Region. Brazzaville: WHO AFRO; 2024. Document AFR/RC74/INF.DOC/7.

18. World Health Organization Regional Office for Africa. Framework for the Implementation of the Immunization Agenda 2030 in the WHO African Region. Endorsed August 2021.

19. World Health Organization, United Nations Children’s Fund. WHO/UNICEF Estimates of National Immunization Coverage (WUENIC), 2025 revision. Geneva: WHO; 2026. Available from: https://immunizationdata.who.int/pages/coverage/wuenic-coverage.html

20. Burton A, Monasch R, Lautenbach B, Gacic-Dobo M, Neill M, Karimov R, et al. WHO and UNICEF estimates of national infant immunization coverage: methods and processes. Bulletin of the World Health Organization. 2009;87:535–541. doi:10.2471/BLT.08.053819

21. Danovaro-Holliday MC, Gacic-Dobo M, Diallo MS, Murphy P, Brown DW. Compliance of WHO and UNICEF estimates of national immunization coverage (WUENIC) with Guidelines for Accurate and Transparent Health Estimates Reporting (GATHER) criteria. Gates Open Research. 2021;5:77. doi:10.12688/gatesopenres.13258.1

22. World Health Organization. Global Health Observatory Data Repository. Geneva: WHO; 2024. Available from: https://ghoapi.azureedge.net/api/

23. Causio FA, Villani L, Mariani M, Pastorino R, De Waure C, Ricciardi W, et al. Vaccination coverage trends in European Union from 1980 to 2020: A joinpoint Regression Analysis. European Journal of Public Health. 2022;32(Suppl 3):ckac129.306. doi:10.1093/eurpub/ckac129.306

24. United Nations, Department of Economic and Social Affairs, Population Division. World Population Prospects 2024 Revision. New York: United Nations; 2024. Available from: https://population.un.org/wpp/

25. World Health Organization. WHO-UNICEF estimates of national immunization coverage: surviving infant populations. Geneva: WHO; 2024. Available from: https://immunizationdata.who.int/pages/coverage/wuenic-coverage.html

26. World Health Organization. WHO/UNICEF estimates of national immunization coverage: Questions and Answers. Geneva: WHO; 2024. Available from: https://www.who.int/news-room/questions-and-answers/item/who-unicef-estimates-of-national-immunization-coverage

27. Mboussou F, Kada S, Danovaro-Holliday MC, Farham B, Gacic-Dobo M, Shearer JC, et al. Status of Routine Immunization Coverage in the World Health Organization African Region Three Years into the COVID-19 Pandemic. Vaccines. 2024;12(2):168. doi:10.3390/vaccines12020168

28. Van Brusselen D, Dubois AH, Bindu LK, Moluh Z, Nzomukunda Y, Liesenborghs L. Not only vaccine hesitancy, but also vaccination campaign hesitancy drives measles epidemics in conflict-torn eastern DR Congo. Conflict and Health. 2024;18(1):14. doi:10.1186/s13031-024-00569-6

29. Bello IM, Lebo E, Shibeshi M, Akpan GU, Chakauya J, Masresha B, et al. Implementation of integrated supportive supervision in the context of coronavirus 19 pandemic: its effects on routine immunization and vaccine preventable diseases indicators in the East and Southern African Countries. Pan African Medical Journal. 2021;38:164. doi:10.11604/pamj.2021.38.164.27349

30. von Elm E, Altman DG, Egger M, Pocock SJ, Gøtzsche PC, Vandenbroucke JP. The Strengthening the Reporting of Observational Studies in Epidemiology (STROBE) statement: guidelines for reporting observational studies. PLoS Medicine. 2007;4(10):e296. doi:10.1371/journal.pmed.0040296

31. Stevens GA, Alkema L, Black RE, Boerma JT, Collins GS, Ezzati M, et al. Guidelines for Accurate and Transparent Health Estimates Reporting: the GATHER statement. The Lancet. 2016;388(10062):e19–e23. doi:10.1016/S0140-6736(16)30388-9

32. KfW Development Bank. Vaccination program funding in cooperation with Gavi, East African Community: Ex-post Evaluation. Frankfurt: KfW; 2023. Available from: https://www.kfw-entwicklungsbank.de/PDF/Evaluierung/Ergebnisse-und-Publikationen/PDF-Dokumente-E-K_EN/EAC_Gavi_2023_E.pdf

33. Masresha B, Luce R, Katsande R, Dosseh A, Tanifum P, Lebo E, et al. The impact of the COVID-19 pandemic on measles surveillance in the World Health Organisation African Region, 2020. Pan African Medical Journal. 2021;39(Suppl 1):192. doi:10.11604/pamj.2021.39.192.29491

